# Association between stress and depressive symptoms and the Covid-19 pandemic

**DOI:** 10.1101/2020.07.28.20163113

**Authors:** Jan S. Novotný, Juan P. Gonzalez-Rivas, Šárka Kunzová, Mária Skladaná, Anna Pospíšilová, Anna Polcrová, Jose R. Medina-Inojosa, Francisco Lopez-Jimenez, Yonas E. Geda, Gorazd B. Stokin

**Author notes:** **Corresponding author**: Gorazd B. Stokin, Translational Neuroscience and Aging Program, Centre for Translational Medicine, International Clinical Research Centre, St. Anne’s University Hospital, Pekařská 53, 656 91 Brno, Czech Republic. **Disclosures**: Authors report no financial relationships with commercial interests.

## Abstract

**Objective:** To date, cross-sectional surveys reported frequency and distribution of mental health disorders on convenience samples impacted by Covid-19. Longitudinal assessment of mental health during Covid-19 in a representative population-based sample, however, is currently largely missing. The aim of this study was to investigate changes in perceived stress levels and depressive symptoms measured before and during Covid-19 pandemic in a representative population-based sample.

**Methods:** Baseline data on stress levels and depressive symptoms from a well-established population-based sample were compared with those obtained from self-administered e-questionnaires distributed during Covid-19 pandemic. A total of 715 participants completed e-questionnaires. Wilcoxon signed-rank test was used to test repeated-measure differences, while between-group differences were analysed using Mann-Whitney and the Kruskal-Wallis tests.

**Results:** Perceived stress levels and depressive symptoms increased 1.4 and 5.5 times, respectively, during the Covid-19 pandemic compared to the time prior Covid-19. Changes in stress and depressive symptoms were most significant in females and did not depend on whether one quarantined alone or with others. Feeling of loneliness during Covid-19 pandemic had the greatest impact on increased stress levels and depressive symptoms.

**Conclusions:** This population-based longitudinal study showed that Covid-19 related measures had significant impact on mental health in a general population with the feeling of loneliness identified as the biggest risk factor. This impact indicates the need of timely and tailored treatment of mental health disorders and integration of preventive mental health measures into global public health policies to protect mental health during future pandemics.

## Introduction

The novel Coronavirus disease 2019 (Covid-19) outbreak in Wuhan, China, evolved rapidly into a pandemic with businesses, governments, and international organizations taking unprecedented actions to limit this threat to global health. At individual level, the Covid-19 pandemic presented several challenges ranging from fear of infection by a poorly understood illness with unclear prognosis to limited possibilities of diagnostics and shortage of personal protection equipment(1). Furthermore, actions to curb the spread of Covid-19 led to implementation of unfamiliar public health measures such as social isolation and distancing, remote education and work, ban of travel and concerns about job security and financial sustainability(2). Several of these measures have already been reported to influence mental health in general population in the past outbreaks(3–5). For example, up to 33% of the surveyed participants reported increased worries during the swine flu in the UK(6), 48% of the general population demonstrated depressive symptoms during Ebola outbreak in Sierra Leone(7), and 57% of subjects reported increased irritability during Severe Acute Respiratory Syndrome (SARS) outbreak in Hong Kong in 2002-2004(8). Although these outbreaks were geographically limited compared with Covid-19, findings from these outbreaks are consistent with those from the earliest studies assessing the impact of Covid-19 on mental health. These studies most commonly estimated the frequency of stress and depression either in specific populations such as healthcare professionals(9–11) or in convenience samples from regions of the world experiencing Covid-19 early in the pandemic such as China(12, 13) and Italy(14). To date, only few studies described the impact of Covid-19 on general psychological distress in time(15–17) with only one containing also pre-Covid-19 data(18). In summary, findings from previous outbreaks, together with the early observations from the Covid-19 pandemic, all support the hypothesis that Covid-19, in particular due to its rapid and global spread with unprecedented governmental actions, causes significant impact on the global mental health. This hypothesis is further corroborated by recent position papers on the impact of Covid-19 on global mental health(19, 20), which noted that large majority of studies to date reported largely cross-sectional data from different convenience samples, which do not critically measure changes in various psychological symptoms in response to Covid-19. Moreover, risk factors and mechanisms underlying possible changes in the psychological symptoms in response to Covid-19 remain poorly understood. To address these gaps, we here took advantage of a well-characterized population-based sample representing randomly selected 1% of the population of the city of Brno, Czech Republic, to critically measure changes in stress and depression during Covid-19 longitudinally and to identify possible risk factors.

## Methods

### Study design and study population

A summary of the baseline examination protocol and general characteristics of the population-based sample has been published previously(21). Briefly, the Kardiovize study, is a prospective longitudinal epidemiological cohort that investigates cardiovascular, neuropsychiatric, and other health-related topics in Central Europe carried out on a representative randomly selected 1% population sample of the residents of Brno, Czech Republic. At the beginning of the Covid-19 outbreak Kardiovize team promptly prepared add-on e-questionnaires to investigate changes in mental health experienced by the Kardiovize study participants during Covid-19 emergency measures in Czech Republic. The inclusion criteria for Covid-19 add-on study were all the participants of the Kardiovize study with available data on stress and depressive symptoms. Those diagnosed with Covid-19 infection were excluded (2 cases). Overview of the recruitment process for the Covid-19 add-on study is shown in eFigure 1 in Supplement. The research protocols of the studies were approved by the Internal Review Board and the St. Anne’s University Hospital ethics committee. All participants of the Kardiovize study as well as of the Covid-19 add-on study signed informed consent.

### Procedure

The Covid-19 add-on study was conducted from April 24 to May 27, 2020. During this time Czech Republic implemented strict public health measures in response to Covid-19, which included national quarantine with closure of schools, shops (except for daily essentials), restaurants and borders, social distancing and obligatory personal protection equipment. Total of 1823 Kardiovize study participants were invited to join the Covid-19 add-on study. E-questionnaire was completed by 715 out of 1823 participants in 4 weeks through an online survey module using validated RedCap software (Research Electronic Data Capture) tool(22). The e-questionnaire consisted of several items, which were assessed during the baseline measurements of the original Kardiovize study, and included also the Perceived Stress Scale(23) (PSS) and Patient Health Questionnaire-9(24) (PHQ-9). The e-questionnaire consisted in demographic characteristics, response to how Covid-19 related government measures affected their daily life and habits, including their experience with quarantine and the use of personal protective equipment, their current medical status (in the Supplement pp 2-5) and psychological questionnaires evaluating stress, depressive symptoms, loneliness, and illness perception.

### Main outcomes

Primary outcomes were stress levels and severity of depressive symptoms. Presence and severity of stress was assessed using PSS, presence and severity of depressive symptoms were assessed using the identical two items of PHQ-9 (before Covid-19) and PHQ-4(25) (during Covid-19) questionnaires. Presence of depressive symptoms was defined as the sum of the score on two PHQ items equal or greater than three(25), stress levels were categorized as low (score of 0-13), medium (score of 14-26), and high (score of 27-40).

Secondary outcomes were age, sex, feeling of loneliness, perceived effect of Covid-19-related measures on daily life, habits and finances, and perception of Covid-19 illness as threatening, and their impact on stress and depressive symptoms. The feeling of loneliness was assessed using UCLA 3-item Loneliness Scale(26), perceived effect of Covid-19 was assessed using new set of self-report questions, and perception of Covid-19 was assessed using Brief-Illness Perception Questionnaire(27) (B-IPQ). Presence of feeling of loneliness was defined as UCLA score equal or greater than six. Item-level analysis was used to assess the perception of Covid-19 measured by B-IPQ.

### Statistical analysis

Descriptive statistics were conducted for the socio-demographic variables and behavioural parameters. McNemar’s test was used to assess differences in prevalence of nominal stress levels and presence of depressive symptoms. Gains of stress and depressive symptoms were calculated as a difference between repeated measures (Covid-19 score minus pre-Covid-19 score).

Missing values were identified in baseline stress (N = 13) and depressive symptoms (N = 19), representing 1.8% and 2.7% of the sample. No missing value imputation was performed, only cases with complete pair of values were used in statistical analysis (702 cases for stress level and 696 cases for severity of depressive symptoms). The missing data were completely at random, non-overlapping in cases, with no observable pattern in their distribution in relation to sex, age, or education. There were no significant differences in mean scores of stress levels and severity of depressive symptoms between participants with and without baseline missing values (see eTable 1 in Supplement). The population included all participants of Kardiovize study who completed the Covid-19 add-on study e-questionnaire. No sample-size calculations were performed.

Normality of the data assessed using Shapiro-Wilk test disclosed violation of the normality rules. As a result, we used Wilcoxon signed-rank test for repeated measure differences between baseline and Covid-19 levels of stress and depressive symptoms. Between-group differences (based on sex, age, loneliness etc.) in gains of stress and depressive symptoms were examined using Mann-Whitney U test and Kruskal-Wallis with Dunn-Bonferroni post-hoc test to correct for multiple comparisons. The respective effect size indicators were calculated and transformed to Pearson’s r for a uniform evaluation of effect sizes.

Data were analysed using SPSS v.21 and figures generated in R v.3.6.3 (https://www.r-project.org/) with ggplot2 (v.1.0.12), reshape2 (v.1.6.4) and pheatmap (v.2.3.3.0) packages. Significance was evaluated at α=0.05 and all testing was 2-sided.

## Results

### Demographic characteristics of the Covid-19 population-based sample

The Covid-19 population-based sample consisted of 715 participants, among whom 336 (47%) were men and 379 (53%) were women, with median age of 46 (range, 24–68; interquartile range, 36–56) (Table 1). Participants were largely well-educated considering majority of them held university degrees (347, 48.7%), followed by those who completed General Certificate of Secondary Education (GCSE) (274, 38.4%). Couples and small families represented approximately half of the population studied.

**TABLE 1.** The basic characteristics of the Covid-19 population-based sample.

|  | Total (N = 715) |  | Males (N = 336) |  | Females (N = 379) |  |
| --- | --- | --- | --- | --- | --- | --- |
|  | N | % | N | % | N | % |
| **Age<sup>a</sup>** | 46 | 36-56 | 44.5 | 36-54 | 48 | 38-57 |
| **Age groups** |  |  |  |  |  |  |
| 24-40 yrs | 265 | 37.1 | 137 | 40.8 | 128 | 33.8 |
| 41-55 yrs | 267 | 37.3 | 123 | 36.6 | 144 | 38.0 |
| 56-68 yrs | 183 | 25.6 | 76 | 22.6 | 107 | 28.2 |
| **Education<sup>b</sup>** |  |  |  |  |  |  |
| Without GCSE | 92 | 12.9 | 46 | 13.7 | 46 | 12.2 |
| With GCSE | 274 | 38.4 | 112 | 33.4 | 162 | 42.9 |
| University | 347 | 48.7 | 177 | 52.8 | 170 | 45.0 |
| **Family members** |  |  |  |  |  |  |
| 1 | 100 | 14.0 | 38 | 11.3 | 62 | 16.4 |
| 2 | 256 | 35.9 | 121 | 36.0 | 135 | 35.7 |
| 3 | 145 | 20.3 | 67 | 19.9 | 78 | 20.6 |
| 4+ | 213 | 29.8 | 110 | 32.8 | 103 | 27.2 |
| **Quarantine spent** |  |  |  |  |  |  |
| with someone | 600 | 83.9 | 288 | 85.7 | 312 | 82.3 |
| alone | 115 | 16.1 | 48 | 14.3 | 67 | 17.7 |
| **Feeling of loneliness<sup>c</sup>** |  |  |  |  |  |  |
| absent | 485 | 67.8 | 245 | 72.9 | 240 | 63.3 |
| present | 230 | 32.2 | 91 | 27.1 | 139 | 36.7 |

Abbreviation: GCSE - General Certificate of Secondary Education
<sup>a</sup>Values presented as median (interquartile range, IQR).
<sup>b</sup>University education includes higher vocational school, bachelor, master, and doctoral degrees.
<sup>c</sup>Presence of feelings of loneliness was defined as score of 6 and more in UCLA 3-item Loneliness Scale.

### General stress levels during Covid-19

We first examined stress that participants may have incurred during the Covid-19 pandemic compared to time prior to Covid-19. Prevalence of moderate to high stress amounted to 253 cases (35.4%, 95% CI, 32.5%–39.7%) and 359 cases (51.1%, 95% CI, 47.4%–54.9%) prior to and during Covid-19, respectively. The number of participants reporting moderate to high stress increased 1.4 times during Covid-19 compared to the time prior to Covid-19 (P<0.001; low stress: odds ratio=3.94, 95% CI=2.79, 5.56; high stress: odds ratio=3.65, 95% CI=0.82, 16.23) (Table 2). More specifically, stress levels increased significantly both in the ratio of individual stress categories, as well as the median of score of the PSS questionnaire (P<0.001) (Table 2 and Figure 1A). This significant increase in stress during Covid-19 was observed in both genders (both P<0.001), however, stress gain was significantly higher in females than in males (P=0.01) (Table 2). Increase of stress during Covid-19 was significant for all age groups (all P<0.001) without between-group differences (P=0.89) (Table 2).

**Figure 1.**
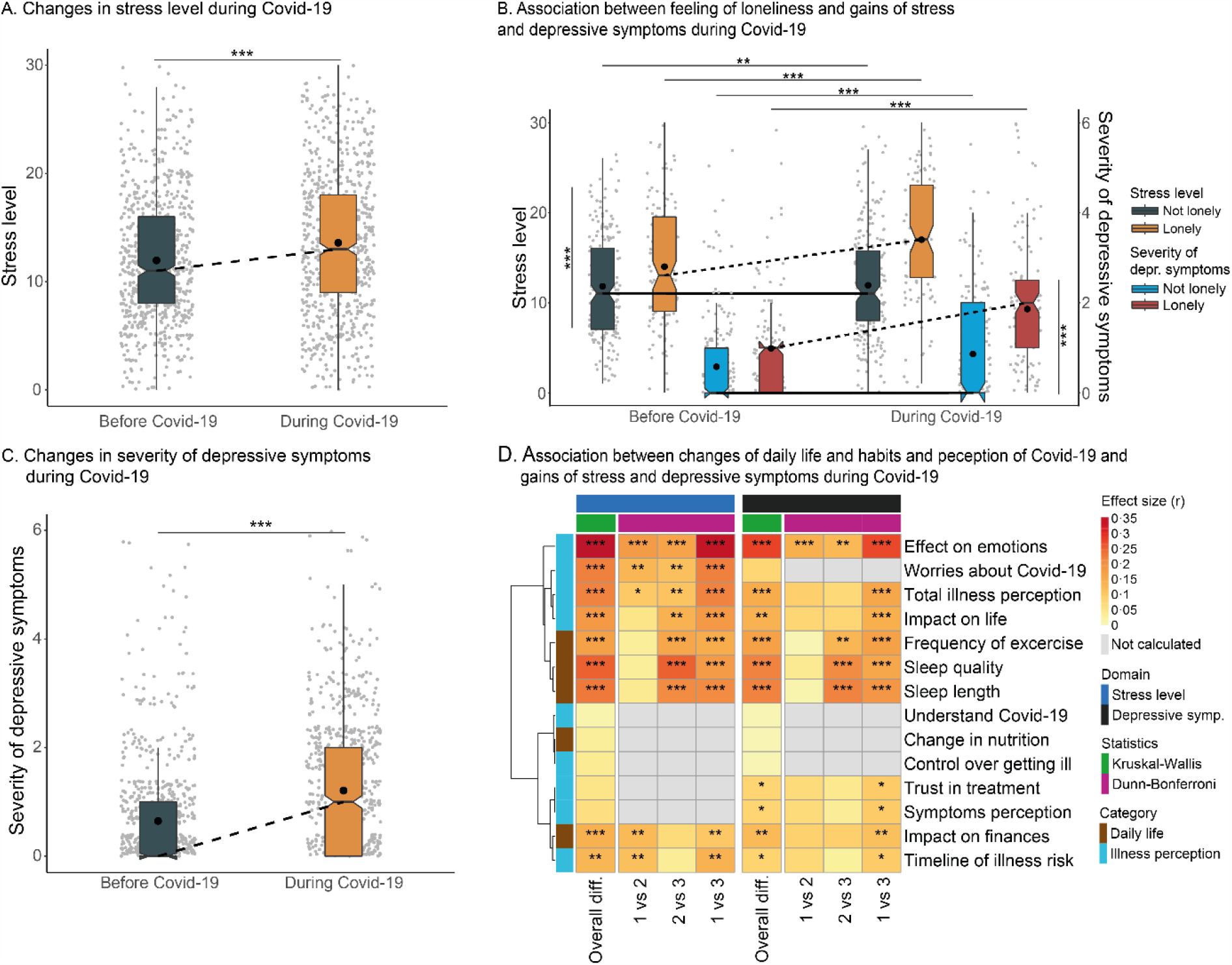
Prevalence of stress level and severity of depressive symptoms before and during Covid-19 and effect of secondary factors on gains of stress level and severity of depressive symptoms^a^. ^a^In A to C, the box plot lines correspond from the bottom of box to the top: 25th percentile, median percentile, and 75th percentile. The whiskers represent the first and third quartiles extended by 1.5 times interquartile range. The dot represents mean, lines indicate changes in medians before and during Covid-19. Horizontal upper bars represent within-group differences with levels of significance, vertical side bar shows between-group difference of gain with levels of significance. In D, clustered heatmap shows Pearson’s r effect sizes (with depicted levels of significance and horizontal clustering using the Wald method) based on Kruskal-Wallis/Dunn-Bonferroni test (with adjustment for multiple comparisons) of effect of individual secondary factors on gains of stress level and severity of depressive symptoms.

**TABLE 2.** Distribution of stress and depressive symptoms before and during Covid-19.

|  | Before Covid-19 |  | During Covid-19 |  | Repeated-measures |  |  | Between-group |  |  |
| --- | --- | --- | --- | --- | --- | --- | --- | --- | --- | --- |
|  | Me | IQR | Me | IQR | TS | P | ES | TS | P | ES |
| **Stress (N = 702)** |  |  |  |  |  |  |  |  |  |  |
| Stress category<sup>a,b</sup> |  |  |  |  | 59.865 | <0.001 |  |  |  |  |
| low | 452 | 64.4% | 348 | 49.6% |  | <0.001 | 3.94 |  |  |  |
| moderate | 241 | 34.3% | 318 | 45.3% |  | 0.12 |  |  |  |  |
| high | 9 | 1.3% | 36 | 5.0% |  | 0.001 | 3.65 |  |  |  |
| Stress level<sup>c,g</sup> | 11 | 8–16 | 14 | 9–19 | -7.33 | <0.001 | 0.28 |  |  |  |
| Stress level by sex<sup>c,d,g</sup> |  |  |  |  |  |  |  |  |  |  |
| males | 11 | 7–14 | 12 | 7–17 | -3.51 | <0.001 | 0.19 | 54589 | 0.01 | 0.10 |
| females | 12 | 8–17 | 15 | 10–20 | -6.60 | <0.001 | 0.34 |  |  |  |
| Stress level by age<sup>c,e,g</sup> |  |  |  |  |  |  |  |  |  |  |
| 24-40 yrs | 12 | 8–17 | 14 | 10–20 | -4.65 | <0.001 | 0.29 | 0.224 | 0.89 |  |
| 41-55 yrs | 12 | 9–17 | 15 | 10–19 | -4.47 | <0.001 | 0.27 |  |  |  |
| 56-68 yrs | 9 | 6–13 | 12 | 6–17 | -3.5 | <0.001 | 0.26 |  |  |  |
| **Depressive symptoms (N = 696)** |  |  |  |  |  |  |  |  |  |  |
| Presence of sign. depressive symptoms<sup>a,f</sup> |  |  |  |  |  |  |  |  |  |  |
| absent | 647 | 93.0% | 427 | 61.4% | 218.85 | <0.001 | 3.90 |  |  |  |
| present | 49 | 7.0% | 269 | 38.6% |  |  |  |  |  |  |
| Severity of depressive symptoms<sup>c,h</sup> | 0 | 0–1 | 1 | 0–2 | -9.32 | <0.001 | 0.35 |  |  |  |
| Depressive symptoms by sex<sup>c,d,h</sup> |  |  |  |  |  |  |  |  |  |  |
| males | 0 | 0–1 | 0 | 0–2 | -4.67 | <0.001 | 0.26 | 52276 | 0.002 | 0.12 |
| females | 0 | 0–1 | 1 | 0–2 | -8.18 | <0.001 | 0.43 |  |  |  |
| Depressive symptoms by age<sup>c,e,h</sup> |  |  |  |  |  |  |  |  |  |  |
| 24-40 yrs | 0 | 0–1 | 1 | 0–2 | -5.08 | <0.001 | 0.32 | 0.007 | 0.99 |  |
| 41-55 yrs | 0 | 0–1 | 1 | 0–2 | -5.73 | <0.001 | 0.35 |  |  |  |
| 56-68 yrs | 0 | 0–1 | 0 | 0–2 | -5.55 | <0.001 | 0.42 |  |  |  |

Abbreviations: Me, median; IQR, Interquartile range; TS, individual tests' statistics; ES, effect size.
<sup>a</sup>No. and percentage (%) are shown for dichotomic and categorical variables.
<sup>b</sup>Repeated-measures were assessed using McNemar-Bowker test ( $\chi^2$ , P) with pairwise McNemar p-value (with Bonferroni correction) and odds ratio effect sizes for pairwise comparisons of nominal variable categories.
<sup>c</sup>Repeated-measures were assessed using Wilcoxon signed-rank test values (Z, P) with Pearson's r effect size.
<sup>d</sup>Between-group differences were assessed using Mann-Whitney U test values (U, P) with Pearson's r effect size.
<sup>e</sup>Between-group differences were assessed using Kruskal-Wallis H test values (H, P) with Pearson's r effect size.
<sup>f</sup>Repeated-measures were assessed using McNemar test results ( $\chi^2$ , P) with odds ratio effect size.
<sup>g</sup>Perceived Stress Scale score with possible range of 0-40.
<sup>h</sup>2-item depressive symptoms score of Patient Health Questionnaire with possible range of 0-6.

### Depressive symptoms during Covid-19

We next examined depressive symptoms before and during Covid-19. Prevalence of depressive symptoms amounted to 49 cases (7%, 95% CI, 5.3%–9.2%) and 269 cases (38.6%, 95% CI, 35.0%–42.4%) prior to and during Covid-19, respectively. The number of participants reporting depressive symptoms increased 5.5 times during Covid-19 compared to the pre-Covid-19 period (P<0.001; odds ratio=3.90, 95% CI=1.89, 8.06) (Table 2). Severity of depressive symptoms also increased significantly during Covid-19 compared to the time prior to Covid-19 (P<0.001) (Table 2 and Figure 1C). This rise in depressive symptoms was present in both sexes (both P<0.001) with females showing higher gain in severity of depressive symptoms than males (P=0.002) (Table 2). Increase of severity of depressive symptoms during Covid-19 was significant for all age groups (median for all age groups 0; all P<0.001) without between-group differences (P=0.99) (Table 2).

### Risk factors for increased stress levels and depressive symptoms

One of the significant features of the quarantine was social isolation and distancing. We therefore asked whether social isolation and distancing play a role in the gain of stress and depressive symptoms. The results indicated that those who spent quarantine alone or with others, showed significant increase of stress levels (both P<0.001) as well as depressive symptoms (both P<0.001) (Table 3). We found no significant difference in gains of stress levels or depressive symptoms between those who spent the quarantine alone or with others (stress level: P=0.77; severity of depressive symptoms: P=0.33).

**TABLE 3.**
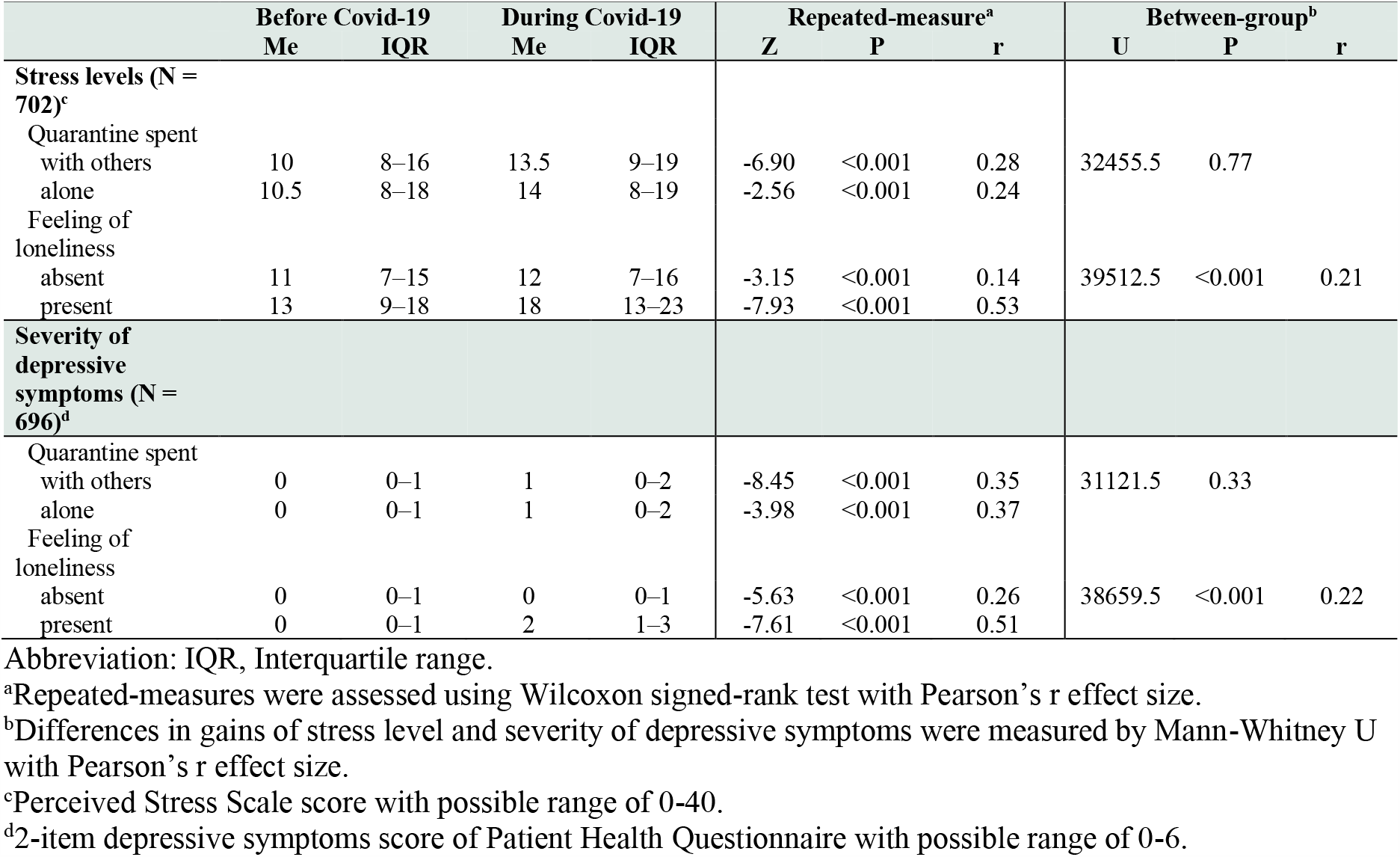
Effect of staying home alone and feeling of loneliness during Covid-19 on gains of stress level and severity of depressive symptoms.

We next investigated whether feeling of loneliness contributes to increased stress levels and severity of depressive symptoms. We observed that participants reporting feelings of loneliness expressed significantly higher gains of stress level (P<0.001), and severity of depressive symptoms (P<0.001). Furthermore, within-group analysis showed that those who reported loneliness exhibited significantly greater increased of stress levels and depressive symptoms (stress level: P<0.001, r=0.53 [95% CI=0.48, 0.58]; severity of depressive symptoms: P<0.001, r= 0.51 [95% CI=0.45, 0.56]) compared with those not feeling lonely (stress level: P<0.001, r=0.14 [95% CI=0.07, 0.21]; severity of depressive symptoms: P<0.001, r= 0.26 [95% CI=0.19, 0.33]) (Table 3 and Figure 1B).

Last, we examined other possible risk factors that might be underlying psychological impact of Covid-19. To this end we assessed perception of Covid-19 disease and changes in daily life functioning. These results showed that the greater gains of stress levels and depressive symptoms were associated mainly with emotional response to the Covid-19 pandemic (Table 4 and Figure 1D). In fact, the strongest association was present for increased negative emotions caused by Covid-19 (P<0.001). Furthermore, increased worry about Covid-19, overall perception of Covid-19 as a threatening disease and worsening of sleep length and quality were found to associate significantly with increased stress levels and severity of depressive symptoms (all P<0.001, see medians in Table 4).

**TABLE 4.**
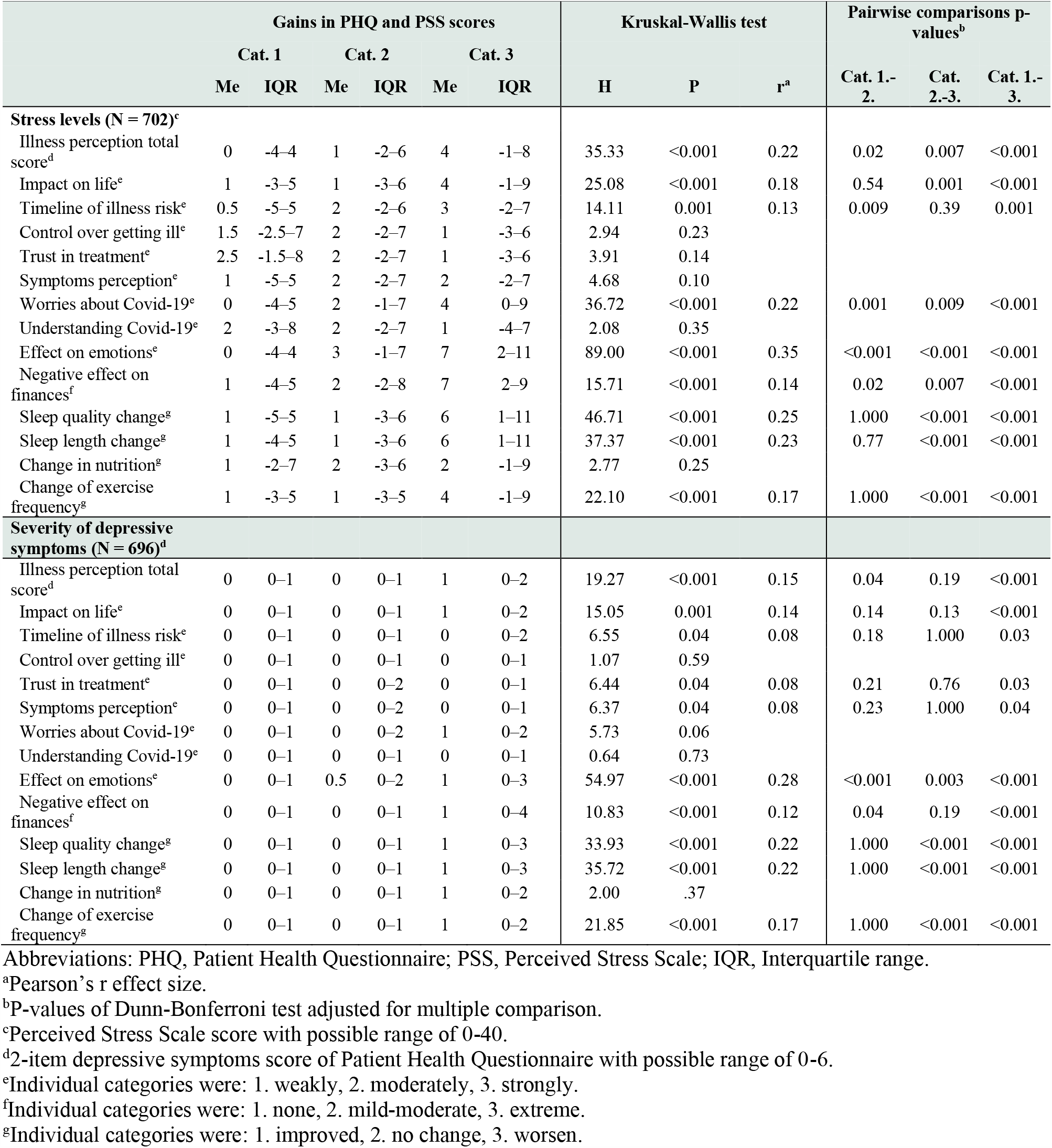
Effect of lifestyle and habits changes, and Covid-19 illness perception on gains of stress level and severity of depressive symptoms.

## Discussions

The main goal of this study was to measure possible changes and magnitude of such changes in stress levels and severity of depressive symptoms before and during Covid-19 pandemic in a representative population-based sample. Our results provide the first longitudinal measure of changes in several mental health indicators following Covid-19. They demonstrate that Covid-19 and related measures led to significant increase in stress levels and depressive symptoms. This increase was consistent across all sex and age groups, revealing higher increase in females compared to males, and no differences among age groups. We find this increase in stress levels and severity of depressive symptoms consistent with cross-sectional(3, 7) and longitudinal(28, 29) data from previous, although largely geographically limited, public health emergencies. The results showed that stress levels increased less than severity of depressive symptoms. This finding can be interpreted by the fact that stress is primarily an acute response to a challenging situation and the participants have already had a few weeks to adapt to the Covid-19 threat and the government imposed Covid-19 measures. Other possible explanation might be that impact of Covid-19 on public health (number of infected, hospitalized and dead) in the Czech Republic was relatively mild, which resulted in lower level of stress. On the contrary, the severity of depressive symptoms has been gradually increasing due to the negative and long-term nature of the Covid-19 pandemic.

There are several explanations for such a significant increase in stress levels and depressive symptoms in response to Covid-19. First, fear of infection by a poorly understood disease of uncertain prognosis and without efficient therapy. Second, implementation of unfamiliar measures to curb the spread of the Covid-19 infection. Third, uncertainty whether governments are managing the Covid-19 pandemic appropriately. Forth, exposure to Covid-19 information and misinformation from media. And fifth, worries about employment and financial stability. Except for the last possible explanation, which has already been documented as a likely cause of stress and depressive symptoms(30–32), there is currently no data about other possible explanations. This strongly suggests that significant effort needs to be made to understand risk factors and other mechanisms underlying increase in stress levels and depressive symptoms in public health emergencies.

The second goal of this study was to identify possible risk factors and underlying mechanisms that play a role in the observed increase in stress levels and depressive symptoms during Covid-19 pandemic. Surprisingly, our results found no association between those who spent Covid-19 period alone or with others. The results, however, disclosed a strong role of emotional processing in the observed Covid-19-related increase in stress levels and depressive symptoms. This manifested in significant effect of negative emotions associated with Covid-19 disease, increased worry regarding the disease, overall perception of Covid-19 as threatening, and extraordinary effect of loneliness. In fact, our data identify feeling of loneliness as the most important risk factor associated with the surge in stress and depressive symptoms. These findings are consistent with previous studies, which established an intimate link between loneliness and psychological distress(33–35).

## Conclusions and recommendations

This study provides the first repeated-measure-based evidence of increase in stress levels and severity of depressive symptoms during Covid-19 compared to the levels prior to the pandemic. Emotional processing of Covid-19 and associated negative feelings played a major role in levels of distress, which was reflected in greater gains of stress and depression in general population with feeling of loneliness identified as the most prominent risk factor. The results show that the exceptional circumstances of Covid-19 pandemic and associated public health measures led to significant negative psychological response in a general population.

Based on these findings, together with bearing in mind the risk of further waves of Covid-19 and likely future outbreaks, public health measures need to integrate efficient strategies to protect mental health. These strategies could include tools for rapid assessment of stress and depression and risk factors such as feeling of loneliness. Identified high risk individuals in particular should have access to appropriate interventions such as enhanced social contact (e.g. in person, via phone, or computer) and cognitive-behavioural techniques(36, 37). Culturally appropriate web-based prevention and intervention tools should also be developed(36, 38, 39). In summary, diagnostics and therapeutics of mental disorders, which increase during pandemic will need to become timely and tailored to the specific measures imposed by the pandemics. In addition, more intense and better organized approaches to mental distress of the general population will need to be integrated into global public health policies to protect mental health during future pandemics.

## Data Availability

The data used in this study are available on request immediately following the publication to anyone who submits the online request that will be approved by the St. Anne University Hospital International Clinical Research Centre internal board. The researches have to provide their research intentions and goals, and specify, and justify requested variables. The data will be provided for a limited and well-defined time via cloud service or e-mail in csv format. After defined period the data should be returned, and all other copies should be destroyed.

## Acknowledgments

The study was funded by the project no. LQ1605 from the National Program of Sustainability II, The Ministry of Education, Youth and Sports Czech Republic (MEYS CR).

## Author Contributions

Drs Stokin and Novotný had full access to all the data in the study and take responsibility for the integrity of the data and the accuracy of the data analysis. The authors contributed to this article as follows: GBS and JPGR conceived the idea of this study, JSN made the statistical analysis, JSN and GBS wrote the draft of the manuscript, all authors contributed to the Critical revision of the manuscript for important intellectual content and reviewed.

## Conflict of Interest Disclosures

Authors report no financial relationships with commercial interests.

## Role of the Funder/Sponsor

The funders had no input in study design, data collection, data analysis, data interpretation, writing of the report, or the decision to submit for publication. All authors had access to study data. The corresponding author had final responsibility for the decision to submit for publication.

## Data sharing statement

The data used in this study are available on request immediately following the publication to anyone who submits the online request that will be approved by the St. Anne’s University Hospital International Clinical Research Centre internal board. The researches have to provide their research intentions and goals, and specify, and justify requested variables. The data will be provided for a limited and well-defined time via cloud service or e-mail in csv format. After defined period the data should be returned, and all other copies should be destroyed.

## Additional Contributions

We are most thankful to Drs. Antonio Pompeiano and Silvie Bělašková for their advice on bioinformatics and statistics. We are grateful to the Translational Aging and Neuroscience Program for fruitful discussions. We thank the participants of the Kardiovize study for their participation on this Covid-19 add-on study.

## Supplement

**Kardiovize Covid-19 e-questionnaire**

1. What is your current weight?
2. How many cigarettes do you smoke per day? If you are a non-smoker, please, fill 0.
3. What is your current family situation?
  a. Living in the relationship with the children
  b. Living in the relationship without the children
  c. Monoparental household (living with one parent)
  d. Living alone
  e. Other (please, specify)
4. How many children do you have?
  a. None
  b. One child
  c. Two children
  d. Three or more children
5. Who are you spending your time with during the quarantine? (multiple choice)
  a. No one
  b. With my partner or spouse
  c. With my children
  d. With other family members
  e. With someone outside my own family
6. During the last 14 days, how often have you been actively and specifically seeking information about the current situation regarding the Covid-19 pandemic and related measures?
  a. Never
  b. Less than once per week
  c. 1-2 times per week
  d. 2-3 times per week
  e. Approximately once per day
  f. Many times per day
7. Does the Covid-19 state of emergency affect your financial situation?
  a. Not at all
  b. Just a little
  c. Pretty much
  d. Extremely
8. How much does the current Covid-19 situation affect your work life? (multiple choice)
  a. The pandemic did not affect my work life/I am currently not working
  b. I have more work than usual
  c. I have less work than usual
  d. I work from home
  e. I changed my job/my job duties or position changed
  f. I stayed home because of kids or family member
  g. I lost my job
9. How many individual PRIVATE (not work-related) social contacts (phone, SMS, Skype, WhatsApp, email, …) have you had in the last 7 days?
  a. None, I am without social contacts
  b. 1 to 3 contacts
  c. 4 −7 contacts
  d. 8 −14 contacts
  e. 15 and more contacts
10. How many individual WORK-RELATED social contacts (phone, SMS, Skype, WhatsApp, email, …) have you had in the last 7 days?
  a. None
  b. 1 to 3 contacts
  c. 4 −7 contacts
  d. 8 −14 contacts
  e. 15 and more contacts
11. Has your sleep quality changed in the past last 14 days?
  a. it got better
  b. it not changed
  c. it got worse
12. Has the length of your sleep changed (on average per day)?
  a. sleep time has increased
  b. sleep time did not changed
  c. sleep time has decreased How often have you exercised in the last 14 days? Write down how many hours per week have you spent performing specific exercises, if zero time, fill in 0.
13. Low intensity exercise (e.g. walking):
14. High intensity exercise (e.g. running):
15. Body building:
16. Stretching:
17. Has the frequency of how often you exercise changed over the last 14 days?
  a. the frequency has increased
  b. the frequency has not changed
  c. the frequency has decreased
18. How many times per week did you go out from your home (work, shop, nature, etc.) in the last 14 days?
  a. Never
  b. 1-2 times per week
  c. 3-5 times per week
  d. Almost every day **How do you follow the government-imposed Covid-19 state of emergency measures?**
19. Are you wearing a mask?
  a. Always
  b. Almost always
  c. Sometimes
  d. Never
20. How often are you washing or disinfecting your hands?
  a. Always
  b. Almost always
  c. Sometimes
  d. Never
21. How often have you respected the restriction of going out?
  a. Always
  b. Almost always
  c. Sometimes
  d. Never
22. How often have you respected the 2-meter social distancing?
  a. Always
  b. Almost always
  c. Sometimes
  d. Never
23. How often have you respected the ban of direct contact with other people?
  a. Always
  b. Almost always
  c. Sometimes
  d. Never
24. How often have you respected the measure that only two people can be in closer contact in public places?
  a. Always
  b. Almost always
  c. Sometimes
  d. Never
25. 25.When do you think the life will get back to the normal in the Czech Republic*? Please indicate the number of months*.
26. How many days did you spend in isolation? *If you have not been in quarantine, please fill in 0*.
  a. because of contact with a person with confirmed coronavirus infection:
  b. because returning from a Covid-19 high risk country:
  c. because you have been tested positive for coronavirus infection:
27. Have you fell ill with Covid-19?
  a. Yes
  b. No
28. What symptoms or signs of Covid-19 have you manifested? (multiple choice)
  c. Fever
  d. Runny nose and cough
  e. Emphysema
  f. Pneumonia
  g. Loss of taste and smell
  h. Headache and dizziness
  i. Nausea, vomiting, diarrhea
  j. Weakness, joints and muscle pain
  k. No symptoms or signs **In the following section we will ask you about your health**.
29. Have you been treated for arterial hypertension?
  a. Yes
  b. No
30. If yes, please provide the name of the medication the and dosage to treat arterial hypertension:
31. Have you been diagnosed with diabetes mellitus type I?
  a. Yes
  b. No
32. If yes, please provide the name of the medication and the dosage to treat diabetes mellitus type I:
33. Have you been diagnosed for diabetes mellitus type II?
  a. Yes
  b. No
34. If yes, please provide the name of the medication and the dosage to treat diabetes mellitus type II:
35. Have you been diagnosed with disease of respiratory-tract (astma bronchiale, CHOPN, etc.)?
  a. Yes
  b. No
36. If yes, please provide the name of the medication and the dosage to treat the respiratory-track disease:
37. Have you been diagnosed with any of the following immune disorders?
  a. Inflammatory bowel disease (e.g. ulcerative colitis or Crohn’s disease)
  b. Rheumatic diseases (e.g. rheumatoid arthritis)
  c. Multiple sclerosis (MS)
  d. Bone marrow transplant
  e. Organ transplant and immunosuppressive therapy
  f. Cancer treated with chemotherapy or radiotherapy
  g. None
38. If yes, please provide the name of the medication and the dosage to treat these disorders:
39. Have you been diagnosed with an allergy or atopic eczema?
  a. Yes
  b. No
40. Are you currently taking medicines containing corticosteroid (e.g. Decamed, Medrol, Depo-Medrol, Dexamethasone, Hydrocortisone, Fortecortin, Methycetone, Fludrocortisone)?
  a. Yes
  b. No
41. Are you currently taking medicines containing Hydrochloroquine (e.g. Plaquenil)?
  a. Yes
  b. No

**eFigure 1:**
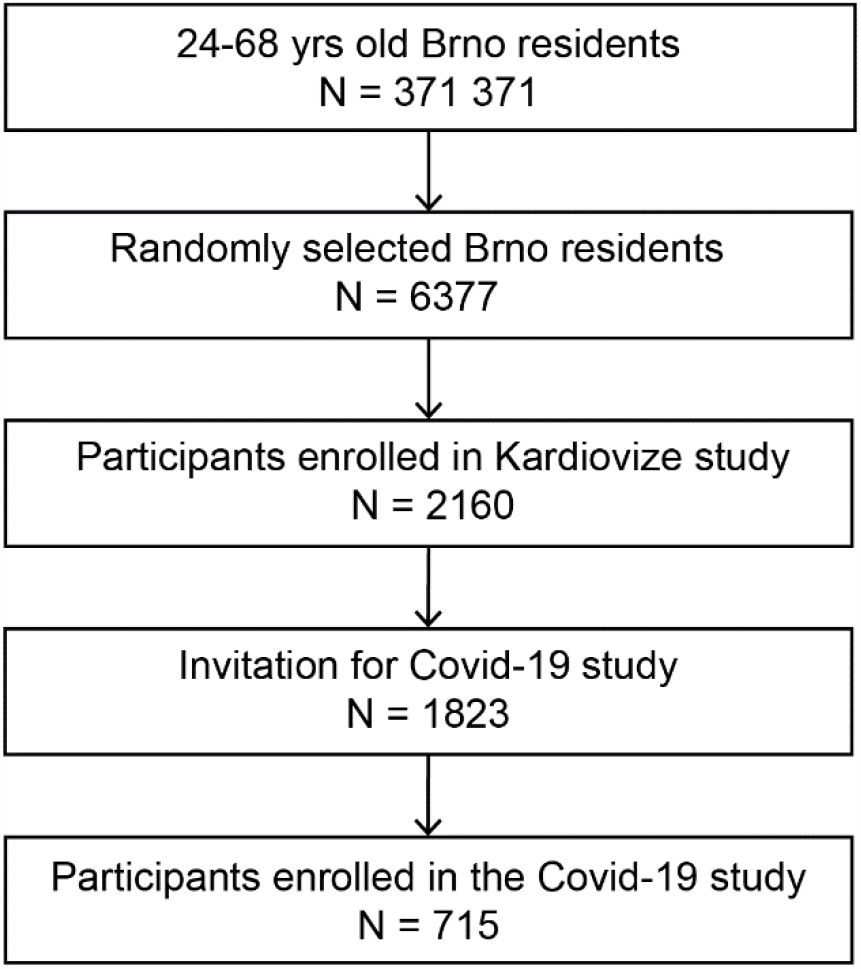
Flow-chart depicting selection of the participants in the Covid-19 research sample.

**eTable 1:**
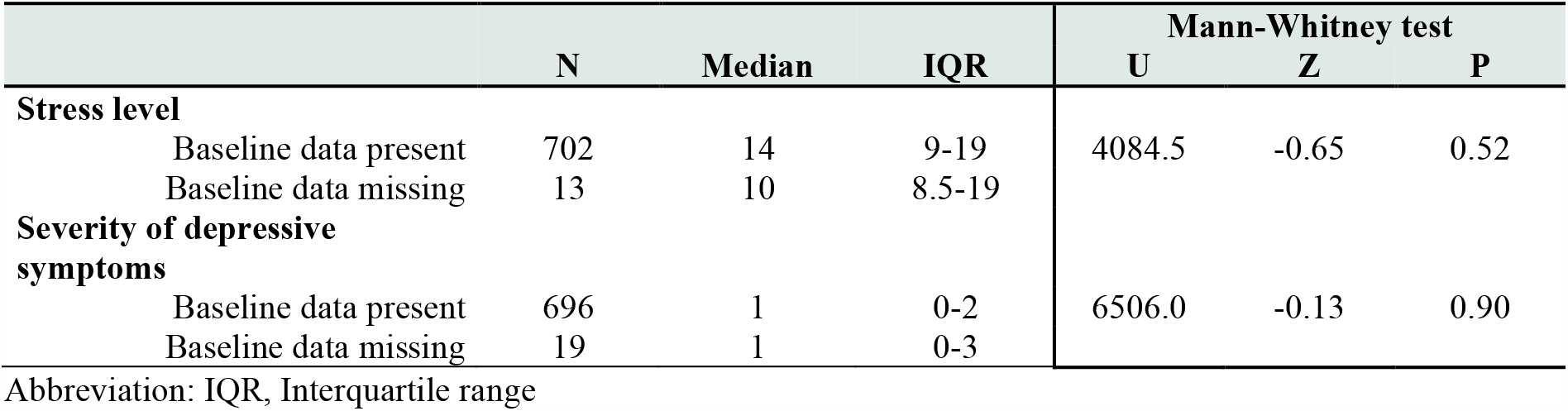
Comparison of Covid-19 stress level and severity of depressive symptom scores between groups with and without baseline missing values.

## Notes

### Competing Interest Statement

The authors have declared no competing interest.

### Author Declarations

The research protocols of the studies were approved by the Internal Review Board and the St. Anne University Hospital ethics committee.

